# Contribution of white matter hyperintensities to ventricular enlargement in older adults

**DOI:** 10.1101/2021.05.11.21256794

**Authors:** Angela CC Jochems, Susana Muñoz Maniega, Maria del C Valdés Hernández, Gayle Barclay, Devasuda Anblagan, Lucia Ballerini, Rozanna Meijboom, Stewart Wiseman, Adele M Taylor, Janie Corley, Francesca M Chappell, Ellen V Backhouse, Michael S Stringer, David Alexander Dickie, Mark E Bastin, Ian J Deary, Simon R Cox, Joanna M Wardlaw

**Author notes:** Corresponding author: Professor Joanna M Wardlaw, University of Edinburgh, Chancellor’s Building, 49 Little France Crescent, Edinburgh, EH16 4SB United Kingdom.

## Abstract

**Background and Purpose:** Ventricular enlargement, especially enlargement of the lateral ventricles, is thought to be positively associated with white matter hyperintensities (WMH). Possible mechanisms behind the association are unclear. Lateral ventricles might increase due to generalised brain tissue loss not specific to periventricular WMH. Alternatively, they may expand into areas of tissue loss related to WMH, take up space and grow in size.

**Methods:** We investigated relations between longitudinal lateral ventricle and WMH volume changes, alongside vascular risk factors, in community-dwelling older people. We assessed lateral ventricle and WMH volumes, accounting for total brain volume, blood pressure, medical assessments and self-reported history of stroke, cardiovascular disease, diabetes and smoking. We used longitudinal data at three time points, each three years apart, between ages 73 to 79, including MRI data from all available time points.

**Results:** Lateral ventricle volume increased steadily with age in all participants, WMH volume change was more variable. Decrease of WMH volume was found in around 20% and increase in remaining subjects. Using a repeated-measurements linear mixed model we found that over 6 years, lateral ventricle volume increased by 3% per year of age, 0.1% per mm Hg increase in mean blood pressure, 3.2% per 1% decrease of total brain volume, and 4.5% per 1% increase of WMH volume. Over time, lateral ventricle volumes were 19% smaller in women than men. No associations were found with other variables.

**Conclusions:** Changes in lateral ventricle volumes and WMH volumes over time are only modestly associated, independent of general brain atrophy.

## Introduction

Enlargement of the lateral and third ventricles occurs in normal aging^1, 2^ and in several diseases including normal pressure hydrocephalus,^3^ multiple sclerosis,^4, 5^ small vessel disease,^6, 7^ mild cognitive impairment, Alzheimer’s disease and vascular dementias.^8, 9^ Commonly, periventricular and deep white matter hyperintensities (WMH) are observed in addition to enlarged ventricles.

In multiple sclerosis, lateral ventricular enlargement appears to be related to periventricular WMH,^4, 5^ as the initially swollen WMH reduce in size over time and are replaced with cerebrospinal fluid. Due to the close proximity of periventricular WMH of presumed vascular origin to the lateral ventricles, it is also possible that the lateral ventricles expand into the areas of tissue loss due to WMH, take up the available space and grow in size.

Most cross-sectional studies that examined ventricular size and WMH of presumed vascular origin found associations between WMH and ventricular enlargement (for full overview of literature see Table I in the Data Supplement). However, it is unclear if WMH of presumed vascular origin, as seen in normal aging, small vessel disease (SVD), mild cognitive impairment, Alzheimer’s disease and vascular dementias,^10^ contribute to the enlargement of the lateral ventricles through direct periventricular tissue damage (as in multiple sclerosis), through shared risk factor associations, or if the ventricles grow because of increased generalised brain tissue loss that is not specific to the tissue affected by the periventricular WMH.

While we know that WMH are associated with vascular risk factors and cerebrovascular disease,^10, 11^ and both severe WMH and large ventricles are associated with an increased dementia risk,^9, 12^ it is not yet clear if WMH and ventricular enlargement share vascular risk factors^13^ and if these vascular risk factors also contribute to ventricular enlargement independently of WMH change. Most studies are cross-sectional with few longitudinal analyses, and many did not account for common covariates including vascular risk factors, or used less sensitive visual ratings (Table I in the Data Supplement). WMH of presumed vascular origin are a common feature of SVD, which is the commonest vascular cause of dementia. Hence, it is important to know if WMH, representative of vascular damage, contribute to ventricular enlargement directly since ventricular enlargement is an important estimate of neurodegenerative changes.

In this study, we investigated the relations between longitudinal changes in lateral ventricle volume and WMH volume, alongside vascular risk factors, over a 6-year period in community-dwelling older people. We hypothesise that increasing WMH contribute to the enlargement of the lateral ventricles, as do vascular risk factors, independent of generalised brain atrophy.

## Materials and methods

We included participants from the Lothian Birth Cohort 1936 (LBC1936), an observational longitudinal cohort study.^14, 15^ All initial participants were born in 1936 and are surviving participants of the Scottish Mental Survey of 1947. At approximately age 70 (Wave 1), 1091 community-dwelling LBC1936 participants agreed to follow-up cognitive, medical and psycho-social assessments. This was followed up by three waves, at mean ages of 73 (Wave 2), 76 (Wave 3) and 79 (Wave 4), where the participants also underwent detailed brain MRI.^16^ We included for this analysis all participants with brain MRI scans at one or more waves. At all waves, participants had blood pressure measurements (mm Hg) and provided self-reported history of stroke (no, yes), history of cardiovascular disease (no, yes), diabetes (no, yes) and smoking (never, ex, current). All participants provided written informed consent under protocols approved by the Scottish Multicentre Research Ethics Committee (MREC/01/0/56) and NHS Lothian Research Ethics Committee (07/MRE00/58).

### Image acquisition

All MRI data were acquired with the same 1.5T GE Signa Horizon HDxt scanner (General Electric, Milwaukee, WI, USA), using a self-shielding gradient set with a maximum gradient strength of 33 mT/m and an 8-channel phased-array head coil. The scanner underwent a detailed quality assurance programme throughout the period of study to ensure scanner stability. Full details of the protocol are described previously.^16^ The MRI acquisition comprised T2 weighted, T2^*^ weighted and fluid-attenuated inversion recovery (FLAIR) scans, acquired in axial plane with a field-of-view of 256×256 mm^2^ and 1×1×2 mm^3^ voxel dimensions (1×1×4 mm^3^ for FLAIR). Also, a 3D whole-brain T1 weighted acquired in the coronal plane with a field-of-view of 129×129mm^2^ and 1×1×1.3 mm^3^ voxel size. The structural sequences remained identical between waves.

### Imaging analysis

All image analysis was performed blind to participant characteristics; additionally the WMH were measured separately and blind to the ventricular measurements (and vice versa) by different trained analysts.

For each participant, all structural magnetic resonance images were registered to the corresponding T2 weighted sequence from the first imaging wave (Wave 2) using FSL-FLIRT.^17^ WMH were defined according to STRIVE criteria.^11^ The intracranial volume (ICV) was semi-automatically segmented in the first imaging wave, as described previously.^16^ We examined changes in ICV between waves. As no changes were found, we used ICV from the first imaging wave to subsequent waves.

For Wave 2, whole brain WMH tissue masks were obtained using a validated semiautomatic segmentation tool.^18^. This method fuses the T2*weighted and FLAIR volumes in red-green colour space to enhance signal differences between tissues and allows the WMH to be separated from other tissues. At Wave 3 and Wave 4 an automatic pipeline was used, the WMH were segmented on the FLAIR images using a modification of a previous method.^19^ WMH were identified by thresholding the raw image intensities to values higher than 1.69 times the standard deviation above the mean intensity of the brain tissue. A lesion distribution probabilistic template^20^ was applied to the thresholded images to exclude hyperintensities unlikely to reflect pathology. Further refinement was achieved by applying Gaussian smoothing, followed by thresholding the smoothed image to remove voxels with an intensity z-score < 0.95 (z-scores were calculated in the raw FLAIR image). Lastly, the intensities below the 10% of the full range in the resultant image were removed. This method reduced the amount of manual editing needed, however all WMH masks were visually checked and if necessary were edited at all three waves. Results from the method used for Wave 3 and Wave 4 are closely correlated to Fazekas scores (Spearman’s ρ= 0.69, 95% CI [0.63 – 0.73], see Figure I in the Data Supplement) consistent with the correlations for the method used at Wave 2.^21^ Stroke lesions (cortical and large subcortical infarcts) were also identified at each wave and excluded from the WMH volume to avoid erroneous measures of WMH change.

To calculate total brain volume we obtained probabilistic masks of white matter, grey matter and cerebrospinal fluid fully automatically from FSL-FAST^22^ using the default parameters. Total brain volume was obtained as the sum of these grey and white matter masks. The Object Extraction Tool in Analyze (AnalyzeDirect, Mayo Clinic, Rochester, Minnesota) was used to extract semi-automatically the lateral ventricles from the 3D T1w volume in all waves, followed by manually editing by a trained analyst.

The WMH volumes, lateral ventricle volumes and total brain volume were recorded in mm^3^ and normalised for the ICV (mm^3^) to account for head size. The volumes of left and right lateral ventricles were combined for a total lateral ventricle volume.

### Statistical analysis

All statistical analyses were performed in RStudio 1.3 with package *lmerTest*.^23^ Plots were made with ggplot.^24^ To observe WMH and lateral ventricle changes we presented them in quintiles in figures. To assess the relations between changes in lateral ventricle volumes and WMH volumes, as well as vascular risk factors, we used a repeated-measurements linear mixed model, fitted by restricted maximum likelihood, with lateral ventricle volume as outcome. The lateral ventricle volume was log transformed to improve normality of residuals in the model. By using a linear mixed model, we could include participants who did not attend all three waves thus maximising use of all available data. The best fitting linear mixed model included a random slope for age, to allow the volume change to vary between individuals over time, and a random intercept for the individual participants, to account for individual differences in lateral ventricle volume. The model was built to include relevant predictors and give the best fit, lowest Bayesian Information Criterion, and residual distribution. We did not include predictors on the grounds of statistical significance but on the basis of relevance for our research question, e.g. brain volume and WMH volume. Other variables were based on available literature (Table I in the Data Supplement) regarding age, sex and relevant vascular risk factors. Since all participants were born in the same year, age was used as a measurement of time. Age was measured in years at the time of imaging and centred to the mean age at Wave 2, to obtain a meaningful intercept in the models. We used the following predictors in the model: age, sex, mean blood pressure, diabetes, history of cardiovascular disease, history of stroke, current smoking status, total brain volume and WMH volume. WMH volume was calculated as the percentage of ICV to prevent a scaling problem. The results were back transformed from log transformation for reporting. Before back transforming, the estimate for total brain volume was divided by 100 for interpretation as a percentage of ICV.

## Results

Over the 6-year period, some participants left the LBC1936^15^ and not all of the remaining participants underwent MRI. Additionally, some participants did not complete MRI due to claustrophobia, or felt unwell at time of MRI. Other participants were excluded because of poor quality scans, e.g. movement, processing problems or missing sequences. Figure 1 summarises imaging data per wave. In total 349 participants provided MRI data at all three waves and the number of participants providing any imaging data at each wave was 675, 482 and 384 respectively.

**Figure 1.**
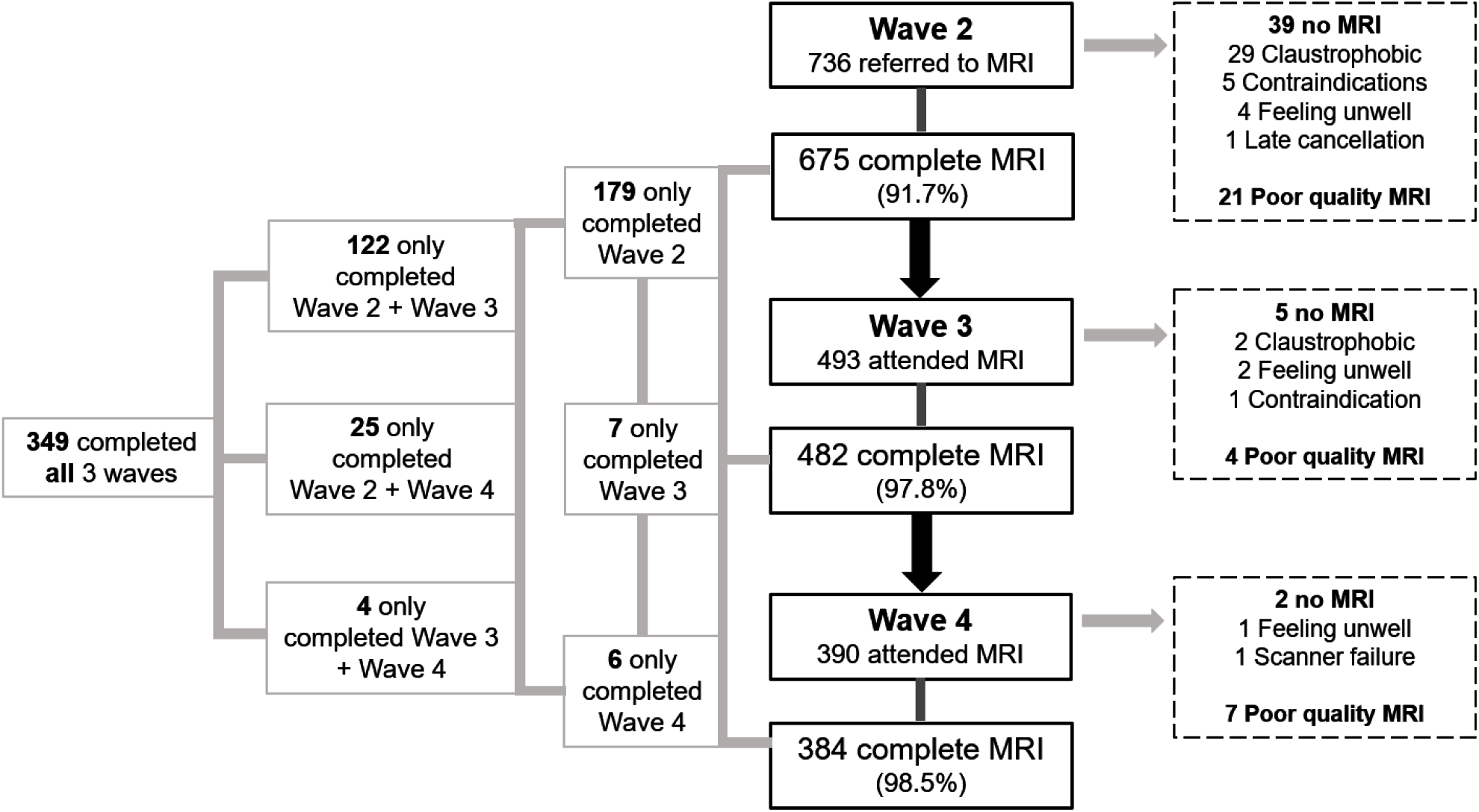
Flow diagram of imaging testing and attrition between waves.

The group of participants who did not complete MRI at Wave 4 had a significantly larger normalised WMH volume at Wave 2 (mean ± SD, 0.009 ± 0.010) than participants who did complete MRI at Wave 4 (0.008 ± 0.008; t(586)= -2.086, p=0.037). They also had a slightly smaller total brain volume at wave 2 (0.678 ± 0.025 vs. 0.685 ± 0.022; t(599)= 3.649, p<0.001), more had diabetes (12.4% vs. 7.9%) and more were current smokers (13.4% vs. 3.3%).

Table 1 shows the demographics for each wave of the study. The proportion of men (∼53%) and women (∼47%) were generally consistent over the three waves. At Wave 4, the mean blood pressure was lower, but the percentage of participants with cardiovascular disease, diabetes and history of stroke was higher. Fewer people were current smokers at Wave 4 compared to Wave 2. The mean lateral ventricle volume increased over time, as did the WMH volume, while total brain volume decreased.

**Table 1.** Demographics per wave of participants with imaging data

|  | **Wave 2**<br>(n=675) | **Wave 3**<br>(n=482) | **Wave 4**<br>(n=384) |
| --- | --- | --- | --- |
| Age, mean (SD) | 72.67 (0.72) | 76.37 (0.65) | 79.44 (0.65) |
| Sex, male (%) | 357 (52.9) | 258 (53.5) | 204 (53.1) |
| Mean blood pressure, mean (SD) | 102.08 (10.89) | 101.55 (11.03) | 99.68 (10.69) |
| Cardiovascular disease, yes (%) | 182 (27.0) | 161 (33.4) | 139 (36.2) |
| Diabetes, yes (%) | 69 (10.2) | 58 (12.0) | 50 (13.0) |
| History of stroke, yes (%) | 46 (6.8) | 53 (11.0) | 53 (13.9) |
| Current smoker, yes (%) | 54 (8.0) | 29 (6.0) | 13 (3.4) |
| Total brain volume*, mean (SD) | 0.682 (0.024) | 0.674 (0.025) | 0.665 (0.024) |
| Lateral ventricle volume*, mean (SD) | 0.022 (0.010) | 0.025 (0.012) | 0.029 (0.013) |
| WMH volume*, mean (SD) | 0.008 (0.009) | 0.010 (0.010) | 0.014 (0.012) |

\* variables expressed as a ratio, normalised for intracranial volume.

### Changes in WMH volume and lateral ventricle volume over time

The changes in WMH volume ratios between waves are visualised in Figure 2, after dividing the sample into quintiles by WMH volume ratio change between two waves, i.e. Wave 2 and 3, Wave 3 and 4, and Wave 2 and 4. Quintile 1 (Q1) represents the smallest change or decrease in WMH volume ratio, and quintile 5 (Q5) the greatest increase. On average, there is WMH volume decrease between all waves in Q1, particularly between waves 2 and 3. In all other quintiles (Q2-Q5) WMH volume shows steady growth but at a different rates, with the most growth in Q5.

**Figure 2.**
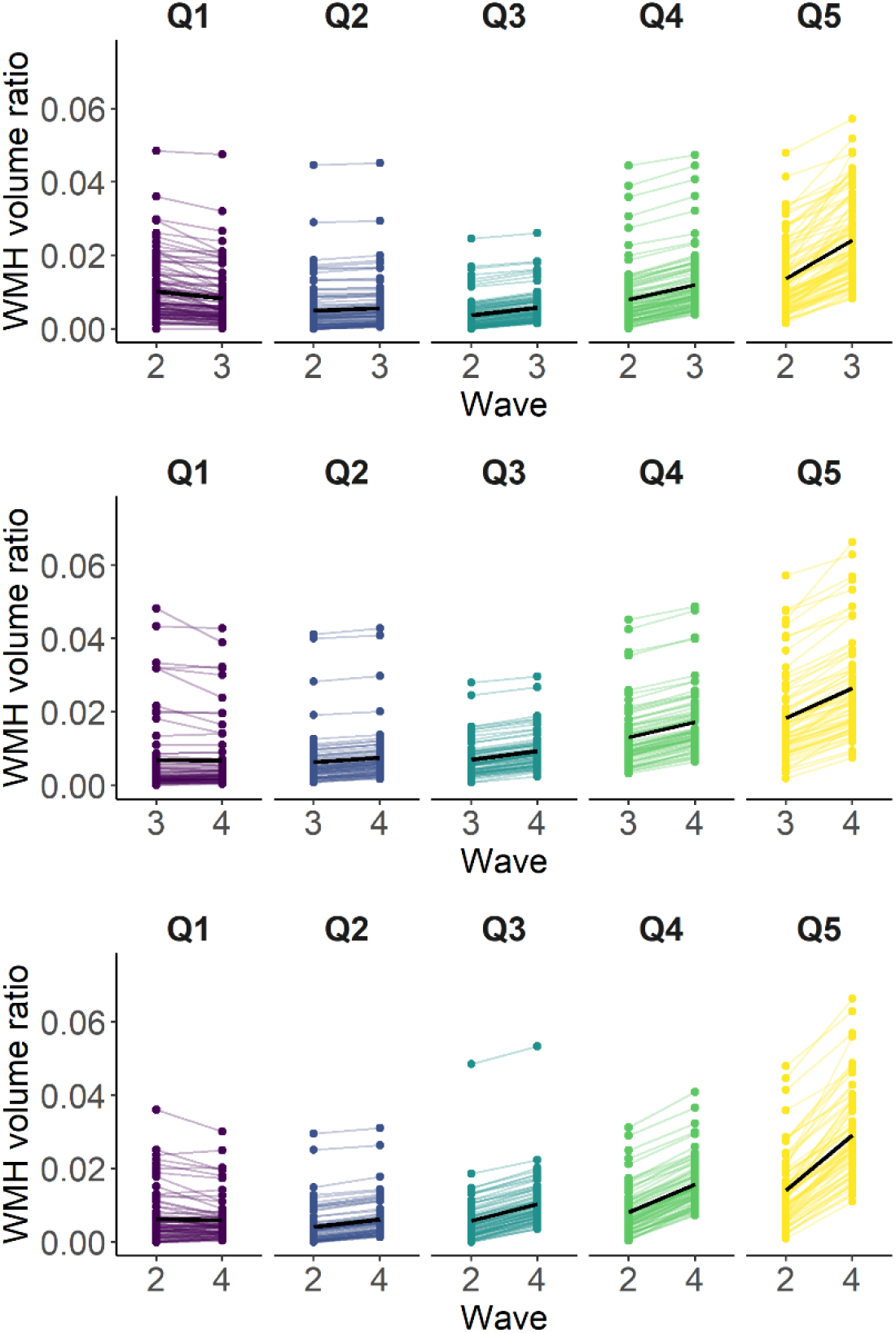
Individual participant’s changes in WMH volume between Waves. Top: Changes in WMH volume between Wave 2 and 3. Middle: Changes in WMH volume between Wave 3 and 4. Bottom: Changes in WMH volume between Wave 2 and 4. The sample is divided by quintiles of WMH volume ratio change between each two waves. Quintile 1 (Q1) indicates no changes or decreased WMH volume ratio; Quintile 5 (Q5) largest WMH volume ratio increase. Black bold lines represent mean volumes per quintile

The changes in volume of the lateral ventricles and WMH over all three waves are shown in Figure 3. The changes are divided per quintile of volume change between Wave 2 and Wave 4, with quintile 1 (Q1) indicating the least increase and quintile 5 (Q5) the biggest increase. Although WMH volume decreased in some participants between Wave 2 and Wave 3 (Figure 2, 3), then showed minor increase between Wave 3 and Wave 4, in contrast, lateral ventricle volumes show a steady increase between all waves in all quintiles, least in Q1 and most in Q5 (Figure 3.

**Figure 3.**
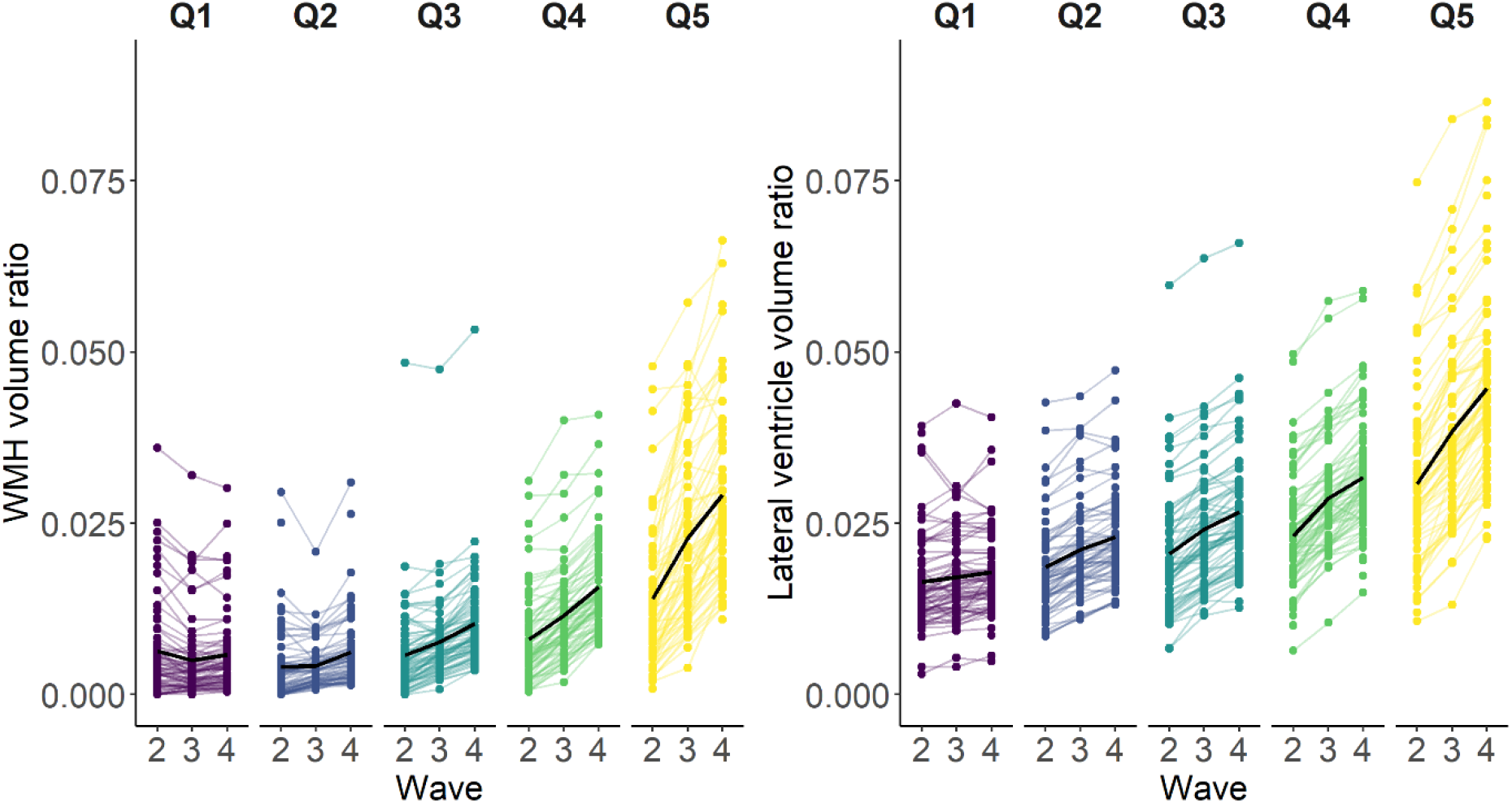
Participant’s WMH volume at all three waves by quintile of WMH volume ratio change between Wave 2 and Wave 4 (Left). Participant’s lateral ventricle volume at all three waves by quintile of total lateral ventricle volume ratio change between Wave 2 and Wave 4 (right). WMH volumes and lateral ventricle volumes are ratios of intracranial volume. Q1 is decrease or smallest increase in and Q5 represents the largest increase. This Figure shows volumes of participants who provided data at Wave 2 and Wave 4 or all three waves.

We then considered the change in lateral ventricular volume according to quintile of change in WMH between Wave 2 and Wave 4 (Figure 4). This showed a wide spread of lateral ventricular volumes in all quintiles (wider spread in Q1 than Q5) but a similar general increase in ventricular volume in each quintile across the three waves.

**Figure 4.**
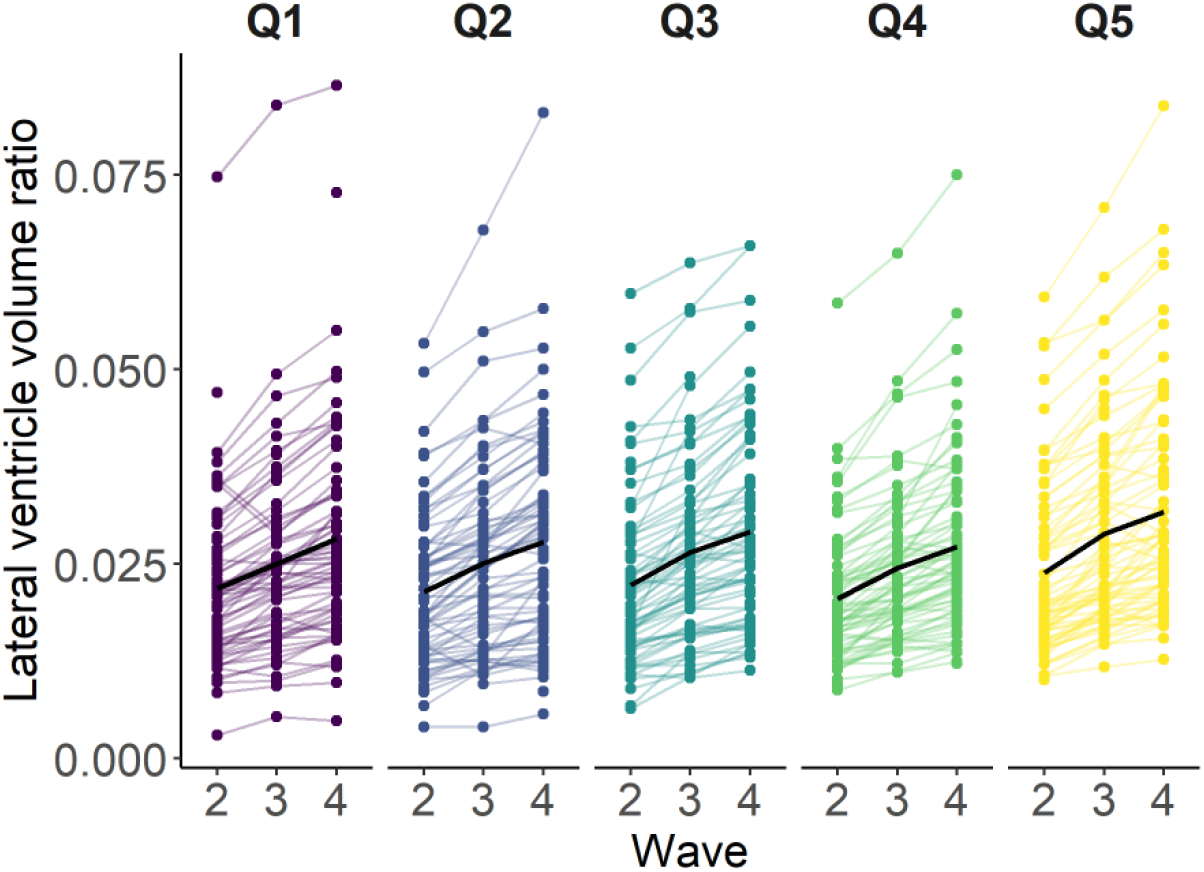
Participant’s total lateral ventricle volume by quintile of WMH volume ratio change between Waves 2 and 4. WMH volumes and lateral ventricle volumes are ratios of intracranial volume. Q1 is decrease or smallest increase in WMH volume ratio and Q5 represents the largest increase in WMH volume ratio. Figure shows volumes of participants who provided data at Wave 2 and Wave 4 or all three waves.

### Repeated-measurements linear mixed model

Table 2 shows the results from the linear mixed model. Since the outcome variable (lateral ventricle volume ratio) was transformed by natural log to improve residual distribution, both the original estimate and the back-transformed estimate are reported. The back-transformed estimates are interpreted as percentage change of the outcome variable per unit change of the predictor.

**Table 2.** Linear mixed model of variables associated with change in total lateral ventricular volume.

| Predictor | Estimate | 95% CI | Exp(estimate) | Exp(95% CI) |
| --- | --- | --- | --- | --- |
| Age<sup>*</sup> | 0.029 | 0.026, 0.032 | 1.030 | 1.026, 1.033 |
| Sex (female) | -0.212 | -0.272, -0.151 | 0.809 | 0.762, 0.860 |
| Mean blood pressure | 0.001 | 0.000, 0.002 | 1.001 | 1.000, 1.002 |
| Diabetes | -0.001 | -0.042, 0.040 | 0.999 | 0.959, 1.041 |
| Cardiovascular disease | 0.010 | -0.013, 0.033 | 1.010 | 0.987, 1.034 |
| History of stroke | 0.025 | -0.009, 0.060 | 1.026 | 0.991, 1.062 |
| Smoking status (ex) | 0.004 | -0.032, 0.040 | 1.004 | 0.968, 1.040 |
| Smoking status (current) | -0.013 | -0.077, 0.050 | 0.987 | 0.926, 1.052 |
| Total brain volume<sup>†</sup> | -3.259<sup>‡</sup> | -3.831, -2.687 | 0.968 | 0.962, 0.974 |
| WMH volume<sup>§</sup> | 0.044 | 0.029, 0.060 | 1.045 | 1.029, 1.062 |

WMH, white matter hyperintensity; Exp, natural exponential function. <sup>\*</sup>centred to mean age in years at Wave 2; <sup>†</sup>normalised for intracranial volume; <sup>‡</sup>estimate divided by 100 before exponentiation; <sup>§</sup>% of intracranial volume. Exp(estimate) is the exponentiated version of Estimate, this is done following log transformation of outcome variable of lateral ventricle volume

Per year of age, the total lateral ventricle volume ratio increased by 3.0%. The total lateral ventricle volume ratio also increased by 4.5% per 1% increase in WMH volume percentage of ICV. The contribution of mean blood pressure was small but significant, as the total lateral ventricle volume ratio increased by 0.1% per mm Hg increase in mean blood pressure. The total lateral ventricle volume ratio is 19.1% lower for women than men, and the total lateral ventricle volume ratio decreases by 3.2% per 1% increase of total brain volume percentage of ICV. History of stroke, cardiovascular disease, diabetes, current or ex-smoking status did not account for variance. As an extra check, we examined if there was an interaction between WMH volume and total brain volume by adding an interaction term to the model, but found no evidence of an interaction (exponentiated estimate; 95% CI: 0.996; 0.678, 1.350; *P*=0.801).

## Discussion and conclusions

In these community-dwelling participants in the eighth decade of life, lateral ventricle volumes increase steadily over a 6-year period, by 3% per year with reference to the intracranial volume. In contrast, WMH volume change was more varied, showing a decrease or little change in some participants while increasing in others. In line with our hypothesis, increasing WMH volume was associated with the enlargement of the lateral ventricles, independent of global brain atrophy. However, the link between WMH and ventricular changes seems small and indirect as shown in Figure 4 where ventricular volume steadily increases regardless of WMH change quintile, and as validated by the modest statistical association. This might suggest that WMH and ventricle volume increase reflect largely indirect and non-overlapping processes.

Other factors that contribute to an increase in lateral ventricle volume are older age and higher mean blood pressure over time. A larger total brain volume may be protective since larger brain volume was associated with smaller lateral ventricles. Females seem to have smaller lateral ventricles than men, even after correction for ICV. This finding is supported by other studies.^2, 9, 25^

Our findings correspond to results from other studies regarding older age^2, 6, 7^ and relation between high blood pressure and (sub) cortical atrophy,^26-28^ although we also account for WMH. Vascular risk factors such as diabetes, history of cardiovascular disease, history of stroke and smoking status did not significantly contribute to change in lateral ventricle volumes. This supports findings in other community dwelling older populations, but also patients with multiple sclerosis or SVD (for an overview see Table I in the Data Supplement). Only one study found an association with prior stroke and cardiovascular risk factors, but used visual assessments for ventricular size rather than volume measurements.^26^

While processes of WMH and lateral ventricle growth do not fully overlap, some of the associations might be explained by the occurrence of demyelination, gliosis and axonal loss immediately adjacent to the ventricles, where WMH often start and progress most rapidly. This damages the tissue, resulting in loss of tissue and more space for ventricles to expand into. One cross-sectional study examined the pathological correlates of WMH and found associations between the total WMH volume, periventricular WMH volume, as seen on MRI, and the severity of the breakdown of the ventricular lining.^7^ A large cohort study found positive correlations between ventricular volume, the lateral and third ventricles, and disproportionate ventricular dilation in healthy older people.^29^ They suggested that the ventricular enlargement is partly caused by SVD-induced atrophy of the periventricular white matter and partly by the active expansion of the ventricles. Another cross-sectional study showed that an increase in WMH contributed to an increase in ventricular volume,^25^ suggesting that changes observed in ventricle size in community dwelling older people and SVD could be related to lost tissue due to white matter damage, as seen as WMH on MRI.

Our findings are supported by a number of strengths. The age range is very narrow as all participants were born in the same year. Since age is strongly related to WMH and ventricular enlargement^13^ (Table I in the Data Supplement), the narrow age range reduces potential confounding effects of age differences. Longitudinal data of people in this age range are rare, in spite of this being a time in life during which there is greater risk of brain structural changes, cognitive decline and dementia. Another strength is the use of volumes of the lateral ventricles, WMH and total brain volume as a measure of atrophy. Visual assessments might be limited by floor and ceiling effects and be less sensitive to longitudinal changes.^30^

Despite these strengths, we examined community dwelling older people who were relatively healthy with only around one third having cardiovascular disease. It is possible that the people with more underlying health issues stopped participating in the study. Participants who were scanned at Wave 2 but not at Wave 4 did have a larger WMH volume at Wave 2 and a smaller total brain volume, which might indicate more atrophy. More of those participants had a history of diabetes and smoked which might suggest they were overall less healthy. However, by using a linear mixed model we did include all available data per wave for every participant, even if some data for that participant was missing, so that all available data contributed to the analysis. Additionally, the balance between participants with and without a history of cardiovascular disease, diabetes, stroke, high blood pressure and smoking was relatively stable over all three waves. Despite using brain tissue volumes to prevent floor and ceiling effects, WMH volumes might not be completely accurate as WMH can be missed or inaccurately identified. We limited these risks by refining the segmentations and a final visual check of the WMH masks.

Further studies should examine ventricular volume in relation to WMH volumes longitudinally. As we also observed decreases and no changes in WMH volume between the first two waves, we would advise to look at changes over a long period of time to identify any decrease in WMH volumes. It would also be interesting to see the relation between ventricular volume and WMH volumes in patients with more severe SVD, stroke or dementias. This might identify the contribution of WMH to ventricular enlargement in brains with larger lesion burden than found in community dwelling subjects.

In conclusion, over a 6-year period we found that, in community-dwelling older people, WMHs contribute to a small proportion of the variance in enlargement of the lateral ventricles, independent of atrophy. However, changes in lateral ventricle volumes and WMH volumes over time were only very modestly linked. Apart from blood pressure, vascular risk factors were not associated with increased lateral ventricle volume. WMH and ventricular enlargement might not share all the same vascular risk factors.

## Data Availability

Data are available upon request to the Lothian Birth Cohort office and Dr Simon Cox. Data request form and more information can be found on the Lothian Birth Cohort website.

## Acknowledgements

The authors thank all participants of the LBC1936 who have contributed, and continue to contribute to the ongoing study. We thank Dr Natalie A. Royle, the radiographers, nurses and other Lothian Birth Cohort 1936 research team members who collected, entered and checked data used in this manuscript.

