## Supplemental Table I for "Contribution of white matter hyperintensities to ventricular enlargement in older adults"

### Abstract

**Background and Purpose.** Ventricular enlargement, especially enlargement of the lateral ventricles, is thought to be positively associated with white matter hyperintensities (WMH). Possible mechanisms behind the association are unclear. Lateral ventricles might increase due to generalised brain tissue loss not specific to periventricular WMH. Alternatively, they may expand into areas of tissue loss related to WMH, take up space and grow in size.

**Methods.** We investigated relations between longitudinal lateral ventricle and WMH volume changes, alongside vascular risk factors, in community-dwelling older people. We assessed lateral ventricle and WMH volumes, accounting for total brain volume, blood pressure, medical assessments and self-reported history of stroke, cardiovascular disease, diabetes and smoking. We used longitudinal data at three time points, each three years apart, between ages 73 to 79, including MRI data from all available time points.

**Results.** Lateral ventricle volume increased steadily with age in all participants, WMH volume change was more variable. Decrease of WMH volume was found in around 20% and increase in remaining subjects. Using a repeated-measurements linear mixed model we found that over 6 years, lateral ventricle volume increased by 3% per year of age, 0.1% per mm Hg increase in mean blood pressure, 3.2% per 1% decrease of total brain volume, and 4.5% per 1% increase of WMH volume. Over time, lateral ventricle volumes were 19% smaller in women than men. No associations were found with other variables.

**Conclusions.** Changes in lateral ventricle volumes and WMH volumes over time are only modestly associated, independent of general brain atrophy.

**Table I.** Overview of literature

| First author | Year | Population (N) | Age, mean ± SD (range) | Study type | WMH | WMH measures | Ventricles | Ventricle measures | Other measures | Covariates | Conclusions |
|---|---|---|---|---|---|---|---|---|---|---|---|
| **Ventricular volume related to WMH** | | | | | | | | | | | |
| COMMUNITY DWELLING ADULTS | | | | | | | | | | | |
| Aribisala[25] | 2013 | Community dwelling elderly (672) | 73±1 | Cross-sectional | Volume ($mm^3$). Absolute values: Median (IQR) | Female: 7,476 (13,838)<br><br>Male: 8,086 (13,842) | Volume ($mm^3$). Lateral, 3rd and 4th ventricles.<br><br>Absolute values: Median (IQR) | Female: 25,052 (14,833)<br><br>Male: 37,010 (20,664) | Total brain volume and GM volumes. Not used in analyses | Factor: **gender.** Covariates: **ICV**, stroke history, diabetes, hypertension, high cholesterol, cardiovascular disease, smoking | WML are associated with brain atrophy, particularly with ventricular enlargement. WML volumes explain 0.9% of the variation in ventricular volume |
| Inatomi[2] | 2008 | Healthy adults (683) | 59 ± 7 (50-88) | Cross-sectional | PVWMH (Fukuda's method; 0-4, normal – diffuse) and DWMH (Fazekas; 0-3) | Incidence PVWMH 40%<br><br>Incidence DWMH 29% | Lateral ventricles; Evans' index | 0.248±0.026 | - | Stepwise regression: **Age**, **sex**, BMI, history hypertension, diabetes, hyperlipidaemia, ischemic heart disease, systolic blood pressure, diastolic blood pressure, haematocrit, HbA1c, total cholesterol, HDL-cholesterol, triglycerides | Ventricular enlargement correlates independently with periventricular white matter changes |
| Palm[29] (thesis) | 2015 | Population study old age (858) | 75.0 ± 5.4 (66-92) | Cross-sectional | Volume (mL). Median | 12.23 | Ventricular volume (mL). Lateral and 3rd ventricles.<br><br>Ventricular dilatation: ventricular | Mean VV = 43.1 | ICV, sulcal CSF volume (total CSF - ventricular CSF) | Age, sex, smoking, hypertension, coronary heart disease, diabetes, BMI, total intracranial volume | WMH volume was positively associated with ventricular volume and ventricular dilatation. WMH volume has a negative association with sulcal CSF volume |

| | | | | | | | | | | | |
|---|---|---|---|---|---|---|---|---|---|---|---|
| | | | | | | | volume (VV) / sulcal CSF volume | | | | |
| West[12] | 2019 | Community cohort (1881) | 62.4 ± 4.5 | Longitudinal 20 years (MRI data only at baseline) | Visual rating of changes in periventricular and deep white matter volume, 1-8 (Barely detectable - extensive changes).<br><br>High grade ≥3 | 223 (12%) with high grade WMH | Visual rating of change, 1-8 (normal - severe atrophy). Ventricles not specified.<br><br>High grade ≥4 | 281 (15%) with high grade ventricular size. | Sulcal size in statistical model 3. | 3 Cox models<br>1: age, sex, race/ethnicity, education APOE status<br>2: model 1 plus smoking status, diabetes, hypertension<br>3: model 2 + MR abnormalities (WMH grade ≥3, ventricular size ≥4, sulcal size ≥3, presence/absence infarcts) | High grade WMH and ventricular size are associated with an increased dementia risk, independent of vascular risk factors |
| NORMAL PRESSURE HYDROCEPHALUS | | | | | | | | | | | |
| Alperin[3] | 2014 | iNPH (8) | (72 – 90) | Case series | Volume (mL)<br>BL 8/8:<br><br>Change 8/8:<br><br>Change 5/8 (clinically improved)<br><br>Change 3/8 | 32.9 ± 37.1<br><br>-5.3 ± 4.0<br><br>-6.1 ± 1.9<br><br>-0.25 ± 0.5 | Volume (mL). Lateral ventricles. BL 8/8:<br><br>Change 8/8: | 127.7 ± 35.1<br><br>-0.5 ± 4.0 | - | - | Gait improved in 5/8 patients who responded positively to ACZ treatment.<br>PVWMH might reflect transependymal CSF |
| MULTIPLE SCLEROSIS | | | | | | | | | | | |
| Dalton[31] | 2002 | CIS (55)<br>Symptomatic (18) | 30.5 (17-49) | Longitudinal 1 year | Number of lesions | - | Volume ($cm^3$). Lateral ventricles. | Median (range) | - | - | Patients with visible lesions (gadolinium) at three months showed significant ventricular enlargement at one year |

| | | | | | | | | | | |
|---|---|---|---|---|---|---|---|---|---|---|
| | | Asymptomatic (37) | 31 (18-50) | | | | BL<br>FU<br>Increase | 5.9 (1.2-39.2)<br>6.3 (1.3-40.6)<br>0.1 (-0.9 – 7.2) | | | |
| Dwyer[4] | 2018 | Total: 192<br>CIS (18)<br>RRMS (126)<br>Progressive (48) | 44.8 ± 11<br>43.8 ± 11.1<br>55.5±7.9 | Longitudinal 5 years | Volume ($mm^3$). BL<br>Atrophied lesion volume<br>Ventricular atrophied volume | CIS: 5,157± 4,427.6<br>RR: 12,170±12,312.0<br>SP/PP: 21,803± 15,108.6<br>CIS: 17.6 ± 17.9<br>RR: 171.1 ± 434.3<br>SP/PP: 298.1 ± 532.4<br>CIS:14.1 ± 17.1<br>RR:139.8 ± 348.1<br>SP/PP: 226.1 ± 439.9 | Lateral ventricles. Volume (mL) Baseline<br>% lateral ventricle volume change | CIS: 27.1 ± 8.7<br>RR: 36.6 ±18.0<br>SP/PP:42.5±18.1<br>CIS: 12.7 (8.7)<br>RR: 16.3 (15.8)<br>SP/PP: 17.4(17.6) | BL normalised brain volume and percent change. Not included in analyses. | Two hierarchical regression models mentioned, not used for analysis. Lesion volumes and ventricular volumes. | Atrophied lesions likely reflect areas that are lost to atrophy and directly replaced by CSF or via atrophy related local movement. Majority of the lesions was periventricular. |
| Sinnecker[5] | 2020 | Total: 127<br>At baseline:<br>CIS (4)<br>RRMS (97)<br>SPMS (22)<br>PPMS (4) | 44 ± 11 (19-66) | Cross-sectional and longitudinal 5 years | Volume (mL)<br>T2w BL<br>T2w FU | 6.1 ± 6.7<br>6.4 ± 7.0 | Lateral ventricles. Volume (mL)<br>BL<br>FU | 29.7 ±14.2<br>33.3 ±16.2 | Deep grey matter atrophy (striatum, thalamus, globus pallidus), normalised brain volume. Whole brain atrophy (estimation). | **Age**, gender | New or enlarging T2w lesions next to the ventricles and thalamic atrophy are associated with enlargement of the lateral ventricles, independent of normalised brain volume. |

| SMALL VESSEL DISEASE | | | | | | | | | | | |
|---|---|---|---|---|---|---|---|---|---|---|---|
| Adamo[32] (not yet peer-reviewed) | 2020 | Total: 166<br>AD (117)<br>Cognitively normal controls (NC; 49) | 71.7 ± 8.5<br><br>69.6 ± 7.6 | Longitudinal 1 year | Volumes ($mm^3$)<br>BL PVWMH<br><br>DWMH<br><br>Growth volume (cc)<br>PVWMH<br><br>DWMH | AD: 6602.2 ± 8076.5<br>NC:3754.7± 5522.9<br><br>AD: 1015.7±1201.1<br>NC:571.1±617.2<br><br>AD: 3.1 ± 3.6<br>NC: 1.6 ± 1.9<br><br>AD: 0.6 ± 0.6<br>NC: 0.4 ± 0.3 | Ventricular CSF (cc)<br>BL<br><br>Growth volumes (cc) | AD: 49 ± 21.5<br>NC: 34.8 ± 17.8<br><br>AD: 8.9 ± 4.9<br>NC: 4.7 ± 1.5 | BL sulcal CSF, supratentorial ICV, brain parenchymal fraction, normal appearing grey matter, normal appearing white matter, right and left hippocampi | **Age, sex, education (years), scan interval,** baseline MMSE, **baseline DRS,** ventricular CSF growth | In AD ventricular CSF growth was associated with PVWMH growth. In NC ventricular CSF growth was associated with both PVWMH and DWMH growth. Ventricular growth might be related to periventricular SVD. |
| Bjerke[33] | 2014 | SVD (46; 34 at FU)<br>BL<br><br>FU | <br><br>74 ± 5<br><br>73 ± 5 | Longitudinal 3 years | Volume ($mm^3$)<br>BL<br>FU | <br><br>24 ± 18<br>22 ± 17 | Dilation (atrophy) 1-8 (no atrophy - severe atrophy). Probably lateral ventricles. | 4.3 ± 1.8 (BL)<br><br>4.1 ± 1.8 (FU) | Ratings sulcal atrophy BL and FU (visual rating). Not used in analyses. | - | WMH volume correlated with ventricular dilation |
| Giubilei[6] | 1997 | VaD (24)<br>Healthy controls (24) | 71.9 ± 6<br>73.8 ± 5.3 | Cross-sectional | Volume (-) | - | Volume (-) Ventricular spaces, not specified | VaD (ratio) 5.1 ± 2<br><br>Controls 3.7 ± 1 | Subarachnoid space volume but not used in analyse. | **Age** | In VaD patients the T2 lesion volumes were related to the increase in volume of the ventricles |

| | | | | | | | | | | | |
|---|---|---|---|---|---|---|---|---|---|---|---|
| Shim[7] | 2015 | Controls (14)<br><br>Systolic hypertension (SH) (11)<br><br>Carotid stenosis (6)<br><br>AD (26) | 84.14±5.99<br><br>84.73±3.29<br><br>72.33±10.69<br><br>74.88±9.18<br><br>At time of clinical evaluation | Cross-sectional<br><br>Pathology (6.79±2.52 years till autopsy) | Total WMH, PVH, DWMH MRI: volumes, normalised by ICV<br><br>Tissue: demyelination (0-3) | WMH: 9.39 ± 10.56<br><br>PVH: 8.30 ± 8.74<br><br>DWMH: 1.09 ± 2.09 | Ventricular volume (mm$^3$, all ventricles; named Ventricular Index, VI)<br><br>Tissue: atrophy of ventricular ependymal (0-3) | 64.43 ± 25.75<br><br>- | - | Sex, years of education, CDR, HIS, hypertension, diabetes<br><br>**Age at clinical evaluation** and duration to death | Volume of WMH, PVWMH, DWMH and the VI increased with age.<br><br>Volume of WMHs and PVWMHs associated with severity of ventricular lining breakdown |
| **Ventricles** | | | | | | | | | | | |
| Apostolova[8] | 2012 | Elderly;<br>NC (46)<br><br>MCI (33)<br><br>AD (43) | 66.4 ±7.8<br><br>73.1 ± 6.0<br><br>75.7 ± 7.6 | Cross-sectional | - | - | Volumes (mm$^3$)<br>Lateral ventricles (frontal, temporal and occipital horns); volume | - | - | **Age**, sex, education | Aging affects the volumes of the hippocampus and lateral ventricles independent of AD pathology |
| Bastin[1] | 2010 | Healthy elderly (90) | 75.7 ± 5.1 (range 68-88) | Cross-sectional | - | - | Lateral ventricles; volumes (mm$^3$) | 1443507 (140390) | ICV | **Age** | Significant correlation between lateral ventricle volume and age after controlling for intracranial volume |
| Carmichael[9] | 2007 | Community dwelling (377)<br><br>Cognitively normal (264)<br><br>MCI (80)<br><br>Dementia (33) | 72.7±3.56<br><br>74.4±4.45<br><br>77.9±5.51 | Longitudinal 4 years | - | - | Ventricle-to –brain ratio (VBR)<br>Volume lateral ventricles/ whole brain (mm$^3$) | - | - | **Age, gender, education level**, presence and incidence of cerebral infarcts, dementia category, **dementia progression** | Lateral ventricles (as measured with ventricle-to-brain ratio) of normal subjects who decline rapidly to dementia are larger than those of normal who remain stable or decline gradually |

| | | | | | | | | | | | |
|---|---|---|---|---|---|---|---|---|---|---|---|
| Manolio[26] | 1994 | Community cohort (303) | 65-95 | Cross-sectional | Visual changes (0-9; no white matter changes – worse than extensive, confluent changes) | (values mentioned per age group for men and women) | Visual assessment (0-9; small-severe enlargement) Probably lateral ventricles | (values mentioned per age group for men and women) | Also visual ratings of sulcal size (sulcal atrophy increased with age) | **Age, sex, prior stroke, hypertension, diabetes, white race** (ventricles) | Atrophy and WMH are associated with age, prior stroke, and known cardiovascular risk factors. |
| Sapkota[34] | 2018 | Total: 723<br><br>AD (439)<br><br>MCI (77)<br><br>VCI (52)<br><br>FTD (125)<br><br>DLB (30) | 70.8±9.4<br><br>71.7±9.3<br><br>70.3±8.2<br><br>71.1±8.2<br><br>66.9±9.1<br><br>73.2±8.8 | Longitudinal 2 years | Volume ($cm^3$) | Total 7.1±10.8<br><br>AD 7.8±11.1<br><br>MCI 3.3±4.8<br><br>VCI 13.2±17.5<br><br>FTD 4.6±6.8<br><br>DLB 7.2±11.6 | Volume ($cm^3$). Ventricular cerebrospinal fluid compartment. Not further specified.<br><br>BL | Total 42.1±20.5<br><br>AD 43.4±19.9<br><br>MCI 31.4±17.2<br><br>VCI 45.6±24.5<br><br>FTD 42.5±21.2<br><br>DLB 42.7±20.0 | Total intracranial volume | Baseline age and apolipoprotein E status | Larger ventricular size at baseline was associated with poorer dementia severity and steeper decline. Association was moderated by early life education and IQ and occupation later in life and is different for sex. Not analysed in relation to WMH. Volumes corrected for ICV |
| Zheng[35] | 2019 | Healthy adults (54) | 43.3 ± 14.9 (21-71) | Cross-sectional | Volume (-) | - | Volume (-) All ventricles | - | Volumes of total grey matter, white matter, deep grey matter nuclei, hippocampi, not used for analyses | **Age** | Volume of WMH, lateral ventricle, inferior lateral ventricle, and 3rd ventricle showed a nonlinear correlation with age |
| **White matter hyperintensities** | | | | | | | | | | | |
| Zivadinov[36] | 2019 | RRMS (176) | 30.7 ± 7.9 | Longitudinal 6 months, yearly for 10 years | Volume (mL)<br>BL<br><br>FU | 7.80 ± 10.50<br><br>11.7±12.8 | Volume (mL) (ventricles not specified) | 41.2±13.6 | Mentions cortical volumes and brain volume, not used in analyses | Age, sex, treatment change | Atrophied T2 lesions were mostly located in periventricular regions and cortical gyri borders |

| | | | | | | | | | | | |
|---|---|---|---|---|---|---|---|---|---|---|---|
| **(Systematic) Reviews** | | | | | | | | | | | |
| Appelman[13] | 2009 | General population<br><br>Cognitive impaired (AD)<br><br>Cerebrovascular risk factors<br><br>Symptomatic vascular disease (Stroke) | - | Cross-sectional<br><br>Cross-sectional<br><br>Cross-sectional<br><br>Longitudinal | - | - | - | - | - | 9/17 studies used covariates. Age, sex, age + sex, age + sex + race or age in combination with vascular risk factors (3/17) | General population: 7/10 studies found relation between WMH and ventricular enlargement of which 1/10 only PVWMH and 1/10 only DWMH.<br>Cognitively impaired: 2/4 found relation between PVWMH and ventricular enlargement, others did not find relation. Several studies also found relation between WML and cortical grey matter atrophy.<br>Cerebrovascular risk factors: 1/2 study found relation between WMH and global atrophy, other study found relation between WMH and ventricular enlargement.<br>Symptomatic vascular disease: 1 study. WMH at baseline correlated with rate of ventricular enlargement. |
| De Guio[37] | 2020 | - | - | - | - | - | Total brain atrophy | - | - | Age, sex, cardiovascular risk factors.<br><br>CADASIL: **age, lacunar volume (1 study)**<br><br>No covariates used in longitudinal studies | Cross-sectional: Lower brain volume + WMH (5 studies), only in upper quartile of WMH. No relationship found in CADASIL patients (4 studies).<br><br>Longitudinal (7): 4/7 reported larger brain atrophy in patients with larger baseline. 3/7 found no relation |

**Bold** covariates are significant. ACZ: acetazolamide; AD: Alzheimer's disease; BL: baseline; BMI: body mass index; CIS: clinically isolated syndrome; CSF: cerebrospinal fluid; diaBP: diastolic blood pressure; DLB: Lewy Body dementia; DRS: Dementia rating scale; DWMH: deep white matter hyperintensities; FTD: frontotemporal dementia; GM: grey matter; HDL: high density lipoprotein; ICV: intracranial volume; iNPH: idiopatic normal pressure hydrocephalus; IQR: interquartile range; MCI: Mild cognitive impairment; NPH: normal pressure hydrocephalus; PPMS: primary progressive multiple sclerosis; PVWMH: periventricular white matter hyperintensities; RRMS: Relapse-Remitting Multiple Sclerosis; SPMS: secondary progressive multiple sclerosis; SVD: small vessel disease; sysBP: systolic blood pressure; VCI: vascular cognitive impairment; VaD: vascular dementia; WMH: white matter hyperintensities; WML: white matter lesions

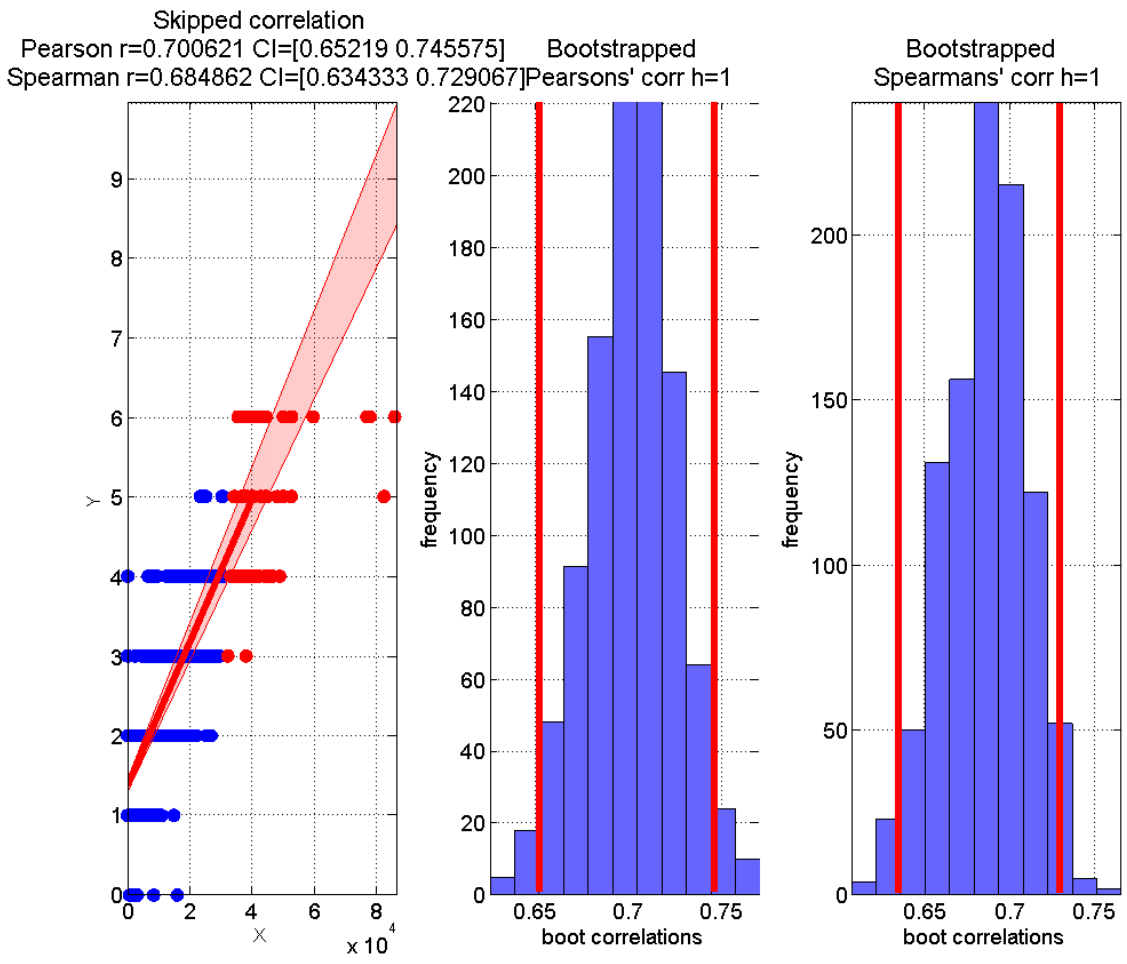


Figure I. Results of skipped correlations between Fazekas scores and WMH segmentation method used at Wave 3 and Wave 4.
